# Sparse Multivariate Functional Cox Landmarking Model with Applications to ALS Progression

**DOI:** 10.1101/2025.06.25.25330277

**Authors:** Andres Arguedas, David Schneck, Annette Xenopoulos-Oddsson, Ximena Arcila-Londono, Christian Lunetta, James Wymer, Nicholas Olney, Kelly Gwathmey, Senda Ajroud-Driss, Ghazala Hayat, Terry Heiman-Patterson, Federica Cerri, Alex Sherman, David Walk, Mark Fiecas, Erjia Cui

**Author notes:** **Correspondence** Andres Arguedas.

## Abstract

Evaluating disease progression over time is crucial for decision-making by patients, clinicians, and health-care providers. Dynamic prediction models update survival curves as new disease progression data become available, often by summarizing the data into scalar values. To reduce data compression, we propose a sparse multivariate functional Cox landmarking model. At each landmark time, our approach first applies sparse multivariate functional principal component analysis (FPCA) to estimate individual disease progression trajectories and then uses a linear functional Cox model to dynamically predict individual survival curves. Simulation studies demonstrate the model’s ability to recover functional coefficients in different scenarios and its superior predictive performance. Our model is applied to Amyotrophic lateral sclerosis (ALS) National History Consortium (NHC) data to dynamically predict risk incorporating the full trajectory of disease progression. External validation using data from Emory University’s ALS Clinic further confirms its predictive advantage over non-functional counterparts.

## 1 INTRODUCTION

### 1.1 Background and Motivation

Evaluating disease progression over time is crucial for assessing patient health and informing clinical decisions. Clinicians often rely on biomarkers and functional measurements as proxies for a patient’s continuously evolving disease state. However, these measurements are often collected at irregular and infrequent clinical visits, leading to sparsely observed data. Such sparse, multivariate data structures highlight the need for specialized methods to effectively capture disease progression. Developing tools that leverage the longitudinal nature of these data is crucial for predicting key outcomes, such as time to death or disease milestones, and ultimately enhancing patient management.

Our work is motivated by the data from patients with amyotrophic lateral sclerosis (ALS), a neurodegenerative disease with no known cure ^1^. Although ALS is quite rare, affecting only 5 out of 100,000 people in the United States, median survival time after diagnosis is short, around 2 years ^2^,^3^. Specifically, the ALS Natural History Consortium (NHC) dataset includes variables measured during routine clinical visits from over 1,000 patients. A detailed description of the ALS NHC dataset is provided in Section 3.1. Among all the measurements, the ALS Functional Rating Scale - Revised (ALSFRSR) ^4^ is one of the most commonly used metric to assess ALS disease progression ^5,6,7,8^. The ALSFRS-R scores are recorded during clinic visits, which may occur sparsely every few months and not at the same time for each individual. Subscores are also defined for the ALSFRS-R, giving more precise information on patients’ abilities for certain domains. Dynamic prediction models offer the possibility to continuously refine the survival probabilities as new ALSFRS-R scores are collected from patients during clinical visits.

### 1.2 Landmarking for Dynamic Prediction

Standard time-to-event models, such as the Cox proportional hazards (PH) model, may incorporate time-varying covariates provided they are measured without error. Because most longitudinal variables do not meet these requirements, additional models have been developed to account for them. The two most common approaches are joint modeling ^9^ and landmarking ^10^. Both approaches allow for dynamic predictions, in which predictions of survival probabilities are updated for individuals as new information becomes available, although the way in which they account for the longitudinal covariates differs radically. When correctly specified, joint models can perform quite well. However, they are subject to assumptions on the random effects and the joint distribution, which can make computations complex and lead to erroneous results when the model is misspecified ^11,12,13^. In contrast, landmark models are easier to fit and impose fewer assumptions, making them suitable for our analysis.

In the landmarking framework a series of landmark times are chosen based on their clinical importance. A summary of the longitudinal covariate up to the landmark time is then used as a covariate in the model. One commonly used summary statistic is last-observation carried-forward (LOCF). More complex summary measures have been proposed, such as those derived from linear mixed models ^14^. Since the value of this summary statistic changes across participants and between models at different landmark times, its relationship with time to death also changes. When using LOCF with few observations over a long time period, landmarking models may lead to biased results. In addition, detailed information about the trajectory of a participant is lost by condensing it into a single summary statistic. An efficient way of utilizing the trajectory of longitudinal data in landmarking models is of special interest.

### 1.3 Functional Data Analysis

Functional data analysis (FDA) ^15^ has become increasingly popular in longitudinal studies. FDA treats data as observations coming from an underlying smooth and continuous function. In practice this underlying function is unobserved, and data are measured with error. Since this function is continuous, functional data are assumed to be infinite-dimensional, which necessitates the use of techniques for high-dimensional data. Special interest lies in the accurate estimation and description of the underlying function and how it relates to covariates and outcomes of interest. When functional data are measured infrequently and at different time points for each individual, they are called sparse functional data. Longitudinal measurements, like the ALSFRS-R scores, are a special case of sparse functional data.

Among all FDA techniques, functional principal components analysis (FPCA) ^16^ extends PCA to functional data, using eigen-functions instead of eigen-vectors to describe the behavior of data without making distributional assumptions. Researchers have extended FPCA to accommodate both univariate and multivariate sparse functional data, using as few as 5 observations on average per individual ^17,18,19^. Recent advances in FDA have also incorporated functional covariates into Cox PH model ^20,21,22^, with additional extensions to dynamic prediction ^23,24,25,26,27,28^. For landmarking models, Yan et al. ^24^, Shi et al. ^25^ and Gomon et al. ^29^ used the FPCA scores directly as covariates instead of the full trajectory. In contrast, Leroux and Crainiceanu ^27^ used the full predicted trajectories from dense functional data as covariates. However, to the best of our knowledge, dynamic prediction models utilizing sparse multivariate functional data through a landmarking approach have not been explored previously.

### 1.4 Two-Step Landmarking Approach

We propose a two-step landmark model that leverages predicted individual trajectories from sparse multivariate functional data to flexibly model time-to-event outcomes. First, the underlying functions generating the observed sparse multivariate data are estimated using sparse multivariate FPCA and predicted individual trajectories for each functional variable of interest are obtained. A functional linear Cox model is then fit using these predicted trajectories to estimate the functional coefficient. This process is repeated for each landmarking time, enabling dynamic prediction of survival probabilities as patient data is accrued. Our model combines the strengths of landmark models and FDA, providing a dynamic prediction model that leverages the full information from participants over time without strong distributional assumptions on their trajectories or its relationship with the time to the outcome.

### 1.5 Organization of the Article

The rest of this article is structured as follows. Section 2 details the landmarking approach and introduces the model. Section 3 presents results from an application of the model to the ALS NHC dataset. Section 4 details a simulation framework and results for the model. Section 5 discusses the main results, possible extensions, and conclusions.

## 2 METHODS

### 2.1 Notation

Suppose there are *i* = 1, …, *N* individuals. Let *T*_*i*_ and *C*_*i*_ indicate their time to death and censoring, respectively. In practice, we only observe *Y*_*i*_ = min(*T*_*i*_, *C*_*i*_) and its associated event indicator variable Δ_*i*_ = *I*(*T*_*i*_ *≤ C*_*i*_), where *I*(*·*) is the indicator function. For each individual, we observe *M* functional variables, denoted *W*_*im*_(*t*_*ij*_) for the *m*-th variable, with *m* = 1, …, *M*. Let *n*_*i*_ denote the number of observations for individual *i*, with *j* = 1, …, *n*_*i*_. The corresponding individual functional domains is *S*_*i*_ = [0, *Y*_*i*_] with *t*_*ij*_ *∈ S*_*i*_. Notice that *t*_*ij*_ depends on *i* and *j*, but not on *m*. These time points need not to be equally spaced within an individual, nor at the same time across individuals. An additional restriction that all *M* functional variables for individual *i* are observed at the same *n*_*i*_ time points was used. Although not a requirement for applying the proposed method, this simplifies notation and corresponds to the data collection procedure from the real world data used.

Assume that *W*_*im*_(*·*) are measured with error from an underlying, unobserved, smooth, continuous functional process denoted as *X*_*im*_(*·*). We can denote *W*_*im*_(*t*_*ij*_) = *X*_*im*_(*t*_*ij*_) + *ε*_*ijm*_, where the *ε*_*ijm*_ are measurement errors, independent across *i, j*, and *m*, such that *E*(*ε*_*ijm*_) = 0 and 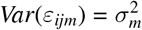 unknown. Thus, *W*_*im*_(*·*) represent sparse multivariate functional data. Additionally, we recorded individual scalar baseline covariates ***Z***_*i*_ for each individual. The observed data for an individual *i* is {*Y*_*i*_, Δ_*i*_, *W*_*im*_(*·*), ***Z***_*i*_}.

Suppose a series of *L* landmark times *s*_*ℓ*_ are chosen, with *ℓ* = 1, …, *L*. Define the corresponding functional domain for each *ℓ* as *S*_*ℓ*_ = [0, *s*_*ℓ*_]. The risk set of individuals at the landmark time *s*_*ℓ*_ is defined as 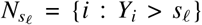, that is, the group of individuals who survived until at least time *s*_*ℓ*_. Consequently, the set of observations from the sparse multivariate functional data which were used are 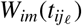, where 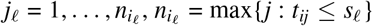, and 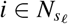.

### 2.2 Multivariate Sparse FPCA

We use the Karhunen-Loève theorem to expand 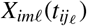 as:

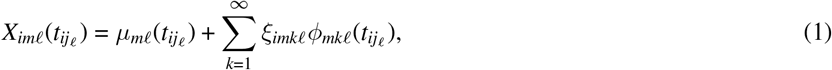

where, for the *m*-th variable, *µ*_*mℓ*_(*·*) is an overall population mean function, *ξ*_*imkℓ*_ are random coefficients that are independent from the 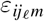, and *ϕ*_*mkℓ*_(*·*) are continuous functions that are orthogonal to each other.

The estimation of *µ*_*mℓ*_(*·*) is done using a P-spline smoother based on all the 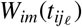, which yields an estimated mean function 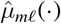. Define the covariance function for the *M* variables across the functional domain as *G*_*ℓ*_, which has elements 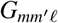, corresponding to the covariance function between the *m*-th and *m*′-th variable. To ensure symmetry of the covariance function with respect to *m* and *m*′, we assume that *m ≤ m*′.

In the case when *m* < *m*′, the estimated smoothed cross-covariance matrix 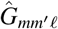 is obtained as the smoothed average using bivariate P-splines of the individual covariances

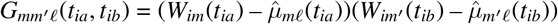

with *a≠ b*. Similarly, when *m* = *m*′, we apply the same procedure to the lower triangle matrix of *G*_*mmℓ*_, thus ensuring the symmetry of the covariance matrix.

The estimated eigen-functions 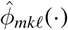 and eigen-values 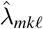 are obtained by solving the following equation:

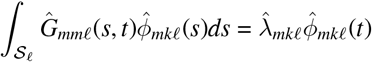

using the constraints that the squared eigen-functions integrate to 1 and they are orthogonal. Using this result, the estimated FPCA scores for participant *i* on the *k*-th eigen-function of the *m*-th variable are approximated as

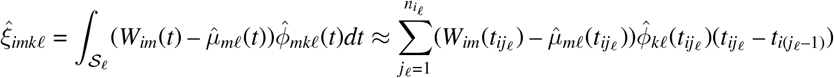

*K* principal components are selected from the infinite number of possible principal components. This is done using the method of cumulative percentage of total variance explained (PVE) based on the estimated eigen-values 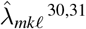 ^30,31^. *A value of 0*.*99 was selected as the cutoff for selecting principal components. Plugging this back into (1), the predicted individual trajectory for participant i* in the *m*-th variable at time *t* becomes:

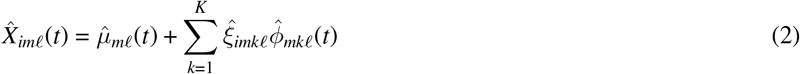

In practice, sparse FPCA can be implemented in R using the `face.sparse()` function from the `face` package ^18^, and the `mface.sparse()` function from the `mfaces` package for its multivariate extension ^19^. Notice that this function not only returns the estimated eigen-functions and eigen-values, but also the predicted mean function and individual trajectories for each participant at time points of interest.

### 2.3 Functional Linear Cox Model

For each landmark time *s*_*ℓ*_, we model the hazard function of death at a certain time *t* after *s*_*ℓ*_ using the following functional linear Cox model:

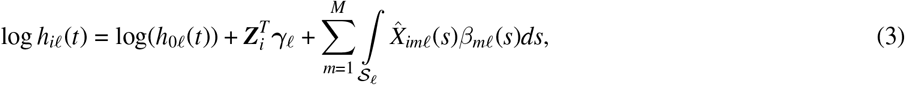

where log(*h*_0*ℓ*_(*·*)) is the baseline hazard function, ***γ***_*ℓ*_ is a vector with scalar coefficients, 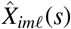 comes from (2), and *β*_*mℓ*_(*·*) are the functional coefficients associated with the *m*-th variable. We can express *β*_*mℓ*_(*·*) using a basis expansion from a spline, say 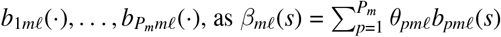, where *θ*_*pmℓ*_ are the spline coefficients. Plugging this in (3) gives:

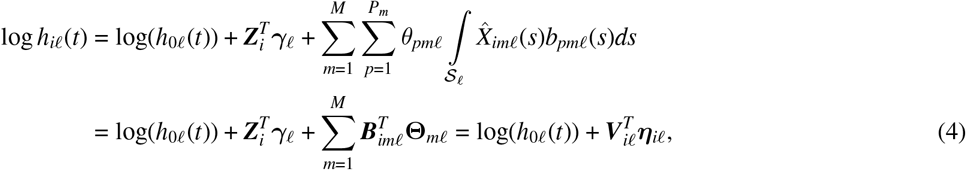

where ***B***_*imℓ*_ is a vector with entries 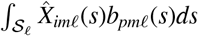, **Θ**_*mℓ*_ is a vector with entries *θ*_*pmℓ*_, which induces 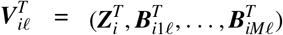 and 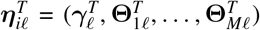. The elements of ***B***_*imℓ*_ were approximated using numerical integration via Riemann sums, using bases formed from cubic regression splines. Based on (4) we can write the penalized partial log-likelihood for the linear functional Cox model as

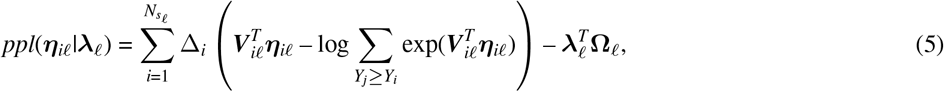

where ***λ***_*ℓ*_ and **Ω**_*ℓ*_ are vectors with elements *λ*_*iℓ*_ and ∥**Θ**_*mℓ*_∥_2_, respectively, with ∥ *·* ∥_2_ denoting the *L*^2^ norm of a vector, which induces a standard quadratic penalty on the *m*-th smooth term **Θ**_*mℓ*_ controlled by a smoothing parameter *λ*_*mℓ*_. For a fixed ***λ***_*ℓ*_ we can minimize (5) using the Newton-Rhapson method to obtain the estimated coefficients as 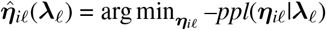.

In practice, the smoothing parameters *λ*_*mℓ*_ are also estimated. Many approaches have been proposed for smoothing parameter selection, such as REML ^32^ or GCV ^33^. Here, we leverage the REML option, which is easy to implement using existing software. Although this modeling approach is complex, it can be fit using existing software such as the `gam()` function from the `mgcv` package; see supplementary material for model implementation details.

## 3 APPLICATION TO THE NHC DATA

### 3.1 The ALS Natural History Consortium dataset

The ALS NHC dataset is a repository of information from people living with ALS from 11 participating multidisciplinary ALS clinics. The ALS NHC dataset contained information from 1,977 participants as of March 2nd 2023. Information collected includes measures of disease progression at each clinical visit, such as the ALS Functional Rating Scale - Revised (ALSFRS-R), as well as baseline participant characteristics. See Berger et al. ^34^ for an overview of baseline characteristics of the cohort.

The ALSFRS-R consists of 12 questions across 4 domains, with questions evaluating a person’s capabilities to perform certain tasks. The 4 domains correspond to: 1) Speech & Swallow (questions 1-3), 2) Fine Motor (questions 4-6), 3) Gross Motor (questions 7-9), and 4) Respiratory Function (questions 10-12). Each question can take values between 0 (indicating severe limitations) and 4 (indicating no limitations at all). A total score and subscores for each domain are calculated by summing the values for each question. The total ALSFRS-R score ranges from 0 to 48, while each of the 4 subscores ranges from 0 to 12, with a lower score indicating more limitations. All four subscores were used as covariates in our analyses. Time to certain disease events of interest are also recorded. The outcome of interest is time-to-death, censoring survival times greater than 10 years to strengthen the clinical relevance of the estimated hazards for all landmark times.

Table 1 presents the main characteristics of the overall sample from the NHC data. The size of the sample was reduced at the 12 month landmark to 800, at 24 months to 554, and at 36 months it was further reduced to 341 participants. One important aspect to highlight is the number of total ALSFRS-R visits, which has a median of 4, with an interquartile range between 3 and 7. This indicates a relatively small number of measured ALSFRS-R subscores.

**TABLE 1.**
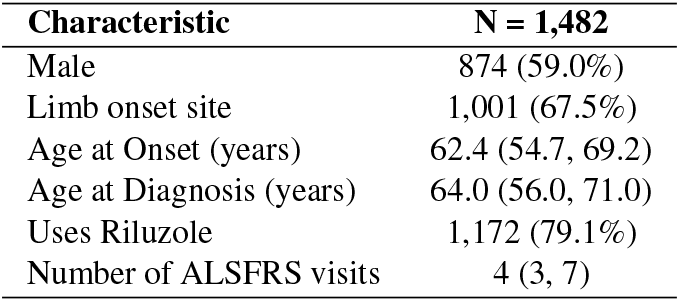
Demographic and clinical characteristics of participants from the NHC data included in the analytic sample. Summary statistics are: median (IQR) for numerical variables and n (%) for categorical variables.

| Characteristic | N = 1,482 |
| --- | --- |
| Male | 874 (59.0%) |
| Limb onset site | 1,001 (67.5%) |
| Age at Onset (years) | 62.4 (54.7, 69.2) |
| Age at Diagnosis (years) | 64.0 (56.0, 71.0) |
| Uses Riluzole | 1,172 (79.1%) |
| Number of ALSFRS visits | 4 (3, 7) |

### 3.2 Multivariate Sparse FPCA

Figure 1 includes the main results from the multivariate sparse FPCA for each of the four subscores. The first two principal components explain over 70% of the variance, while the first five around 98%. The first eigen-function is close to zero for the Speech & Swallow subscores, but not for the others, which allows Speech & Swallow to be separated from them. For the second eigen-function, it has similar trajectories close to zero for both the Fine and Gross Motor subscores, while having larger values for the other subscores. This separates the Fine and Gross Motor subscores, which are similar, to the Speech & Swallow and Respiratory Function subscores. Although more complex the other eigen-functions are interpreted in a similar manner.

**FIGURE 1.**
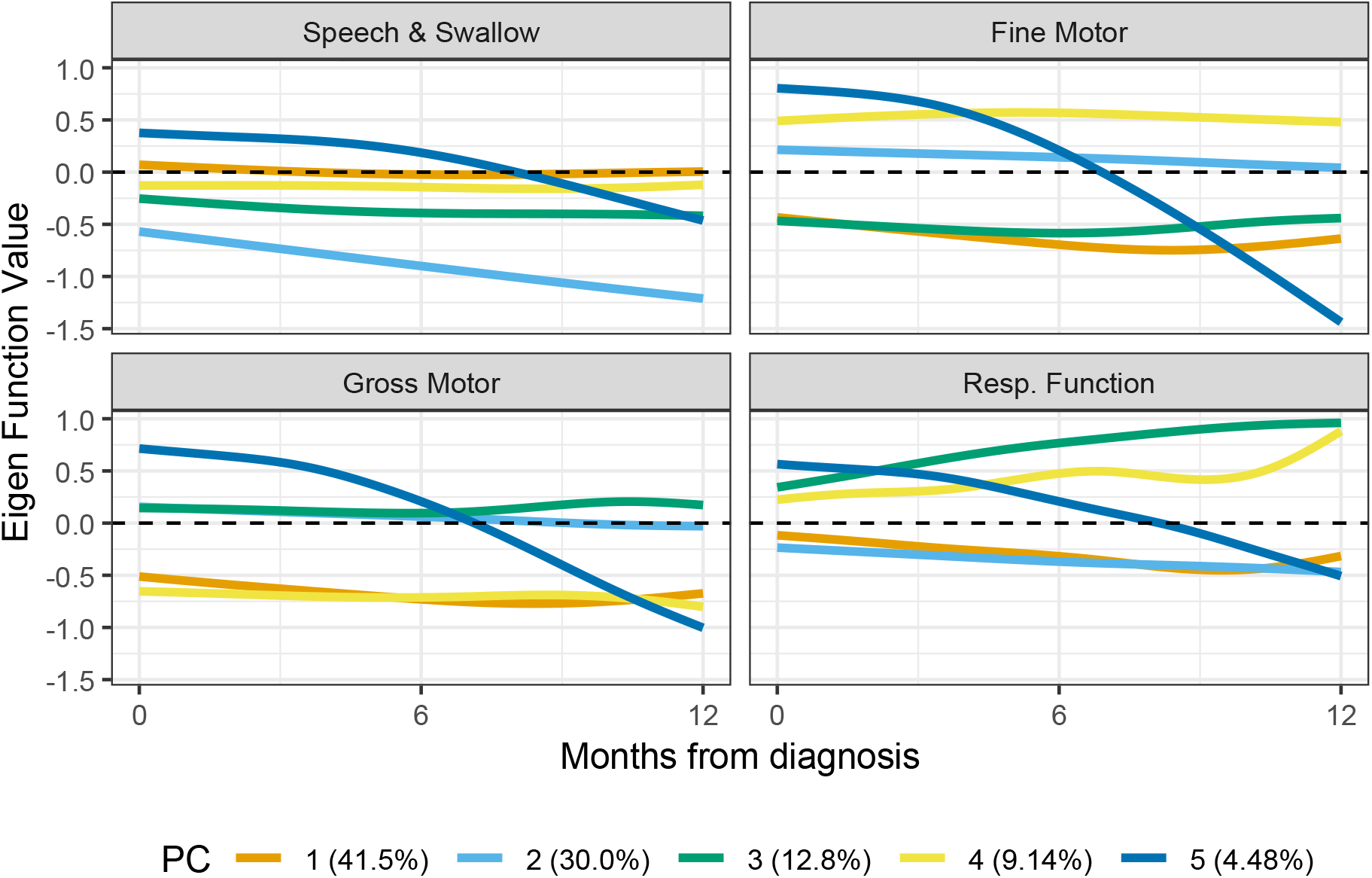
First five estimated eigen-functions from the multivariate sparse FPCA, with their corresponding percentage of variance explained, for each of the ALSFRS-R subscores

Figure 2 presents the main results from the FPCA. These include the estimated mean and variance functions, with corresponding 95% confidence band for the mean function, as well as the estimated correlation matrix, for all four ALSFRS-R subscores.

**FIGURE 2.**
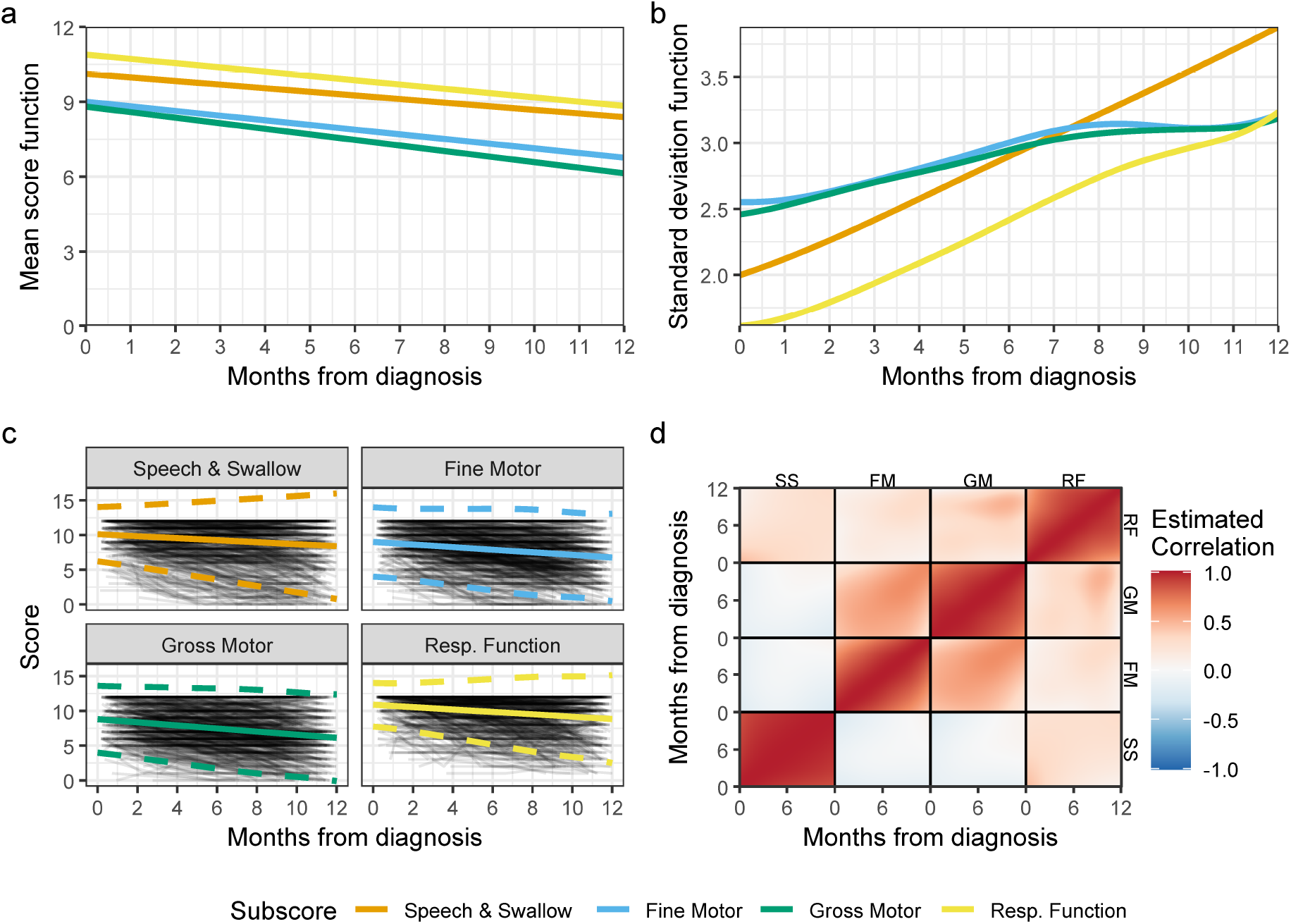
Results from the multivariate sparse FPCA applied to the ALSFRS-R subscores at the one year landmark. **(a)** Estimated mean function. **(b)** Estimated standard deviation function. **(c)** Estimated mean function with 95% confidence bands. **(d)** Estimated smoothed correlation matrix. Abbreviation FM corresponds to Fine Motor, GM to Gross Motor, RF to Respiratory Function, and SS to the Speech & Swallow subscores, respectively

From Figure 2a, the overall population trend is decreasing as expected. The mean function for each subscore is different, with the Respiratory Function subscore having the highest values, while the Gross Motor subscore has the smallest ones. The Speech & Swallow subscore has the smallest slope, indicating less overall change over time compared with the other subscores. A comparison between this estimation and the individual patient trajectories is presented in Figure 2c. The estimated mean function strikes a balance between participants who have high values, and are relatively consistent across time, and those who show a steady decline over the first year. The 95% confidence band for these estimates grows as time goes on, while the rate of growth changes between the different subscores. The estimated standard deviation functions from Figure 2b reflect this. The standard deviation function for the Fine and Gross Motor subscores have a much slower increase and stagnate after 8 months, while both the Respiratory Function and Speech & Swallow subscores have consistently increasing standard deviation functions.

The estimated correlation matrix is presented in Figure 2d. The correlation within subscores is positive and high. Participants with high scores tend to show slower declines than those with lower scores at baseline. The multivariate sparse FPCA gives us an additional estimate of the correlation between subscores. We can notice a relatively high correlation over time between the Fine and Gross motor subscores, while the Respiratory Function and Speech & Swallow subscores show much smaller correlations with all other subscores. The Speech & Swallow subscore is negatively correlated with both the Fine and Gross motor subscores.

### 3.3 Functional Cox Landmarking Model

Having implemented the sparse multivariate FPCA, we obtain the predicted individual trajectories 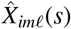 for each of the subscores. A functional Cox landmarking model (FCLM) was fit for the first landmarking time of 12 months after diagnosis. The estimated functional coefficients for each of the four ALSFRS-R subscores from this model are presented in Figure 3.

**FIGURE 3.**
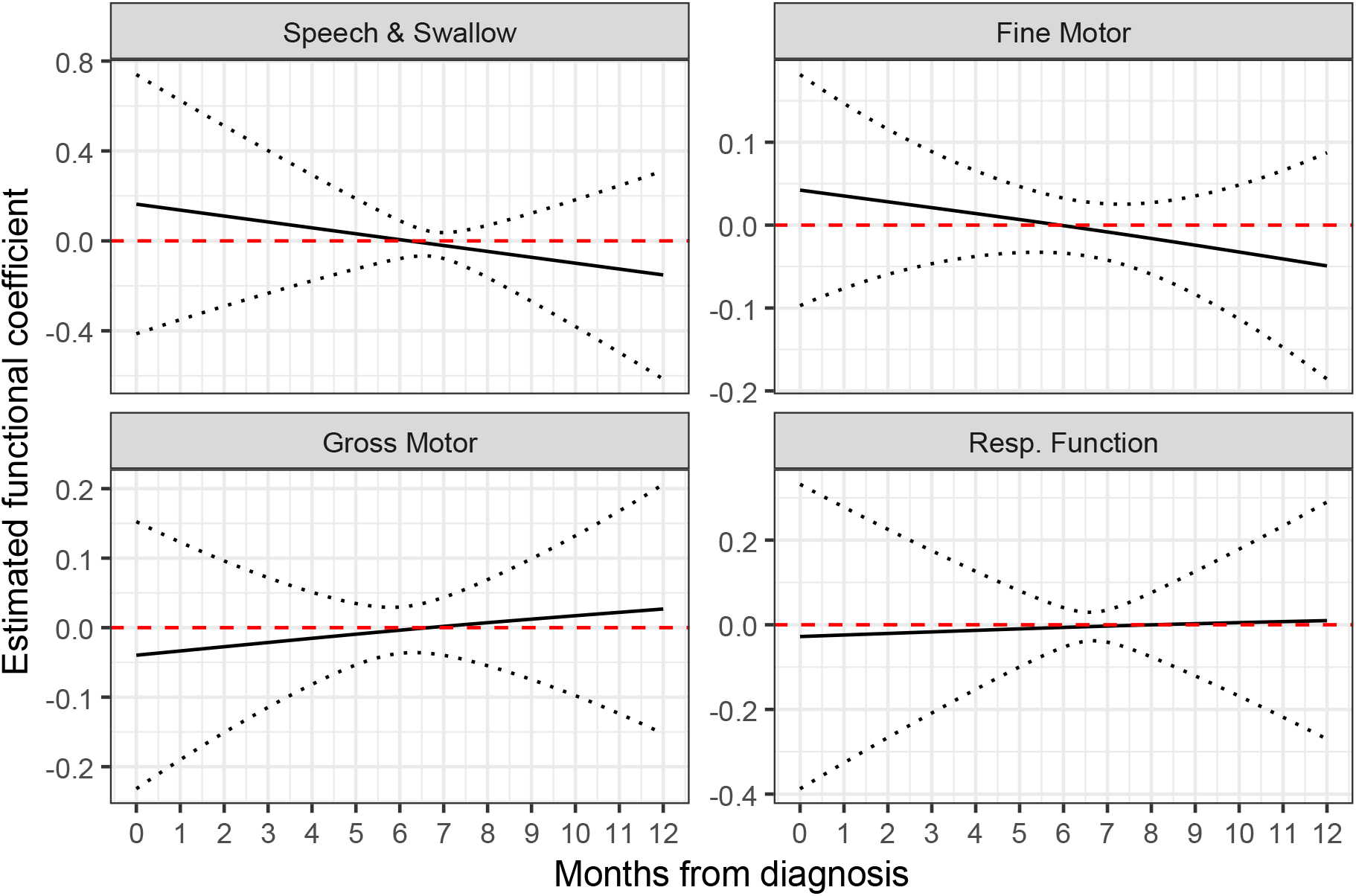
Estimated functional coefficient 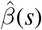 with 95% confidence band, for each of the ALSFRS-R subscores, across time, at the one year landmark

The fact that the ALSFRS-R subscores are expected to be non-increasing gives sense to the interpretation of these functional coefficients. Suppose participants A and B have the same Speech & Swallow subscore trajectories across the first 12 months, but A shows a higher value at diagnosis. In this case, we would expect A’s survival probability to be lower, as the steeper decline in their subscore suggests faster disease progression compared to B. Conversely, if A and B only differ in their Speech & Swallow subscores at the final assessment at 12 months, the participant with the sharper decline would be considered to have a faster progression, indicating higher risk. Therefore, participants with consistently high Speech & Swallow subscores are expected to have the lowest risk. The functional coefficients for the other subscores can be interpreted in a similar manner. The confidence bands for the functional coefficients were obtained using the estimates of the standard error from `mgcv` directly. Bootstrap confidence bands could be preferred to give more accurate results compared with the seemingly large variability from the model fit using `mgcv`.

Landmarking times of 24 and 36 months after diagnosis were used to fit additional FCLMs. Web Figures 1 and 2 present the functional coefficients for these models. The results from the predicted survival probabilities from the models using the three landmarking times are presented in Figure 4.

**FIGURE 4.**
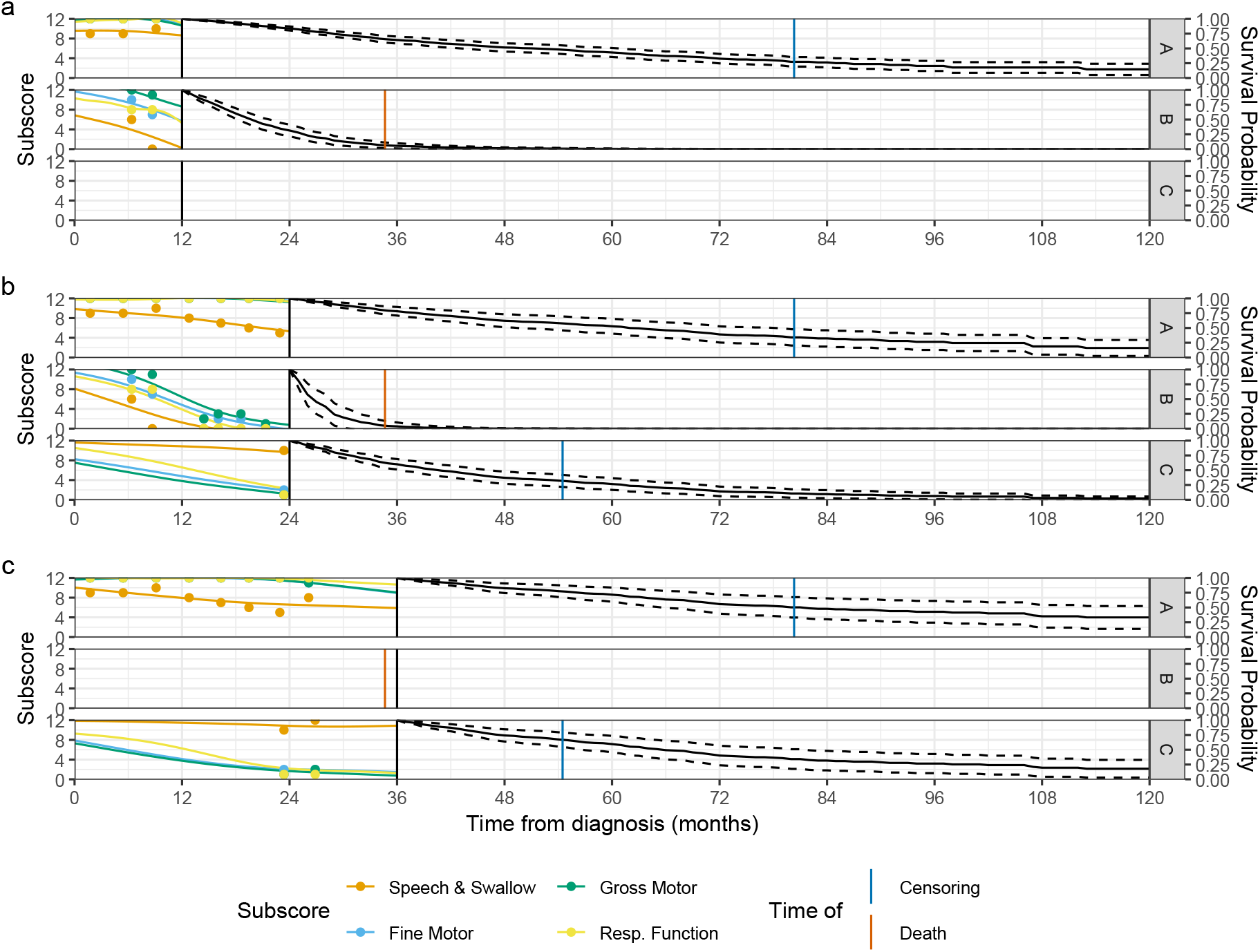
Individual predicted trajectories from the multivariate sparse FPCA for each subscore, as well as the observed ALSFRS-R subscores, and predicted survival probabilities, at the **(a)** 12 **(b)** 24 and **(c)** 36 months landmarking times

We can see the change in the observed subscores across time, as well as the corresponding estimated predicted individual trajectories, which are used to dynamically predict the survival curves for individuals as more disease progression data is accrued. Comparing participants between Figures 4a and 4b, participant A seems to have stable scores over time. Thus their predicted survival probabilities are similar across landmark times. In contrast, participant B has a sharp decrease in their subscores after 12 months, which is reflected by a steep drop in their survival probability at the 24 month landmark time in Figure 4b. For participant C, no information about their ALSFRS-R was collected before 12 months, so personalized trajectories can not be obtained for Figure 4a. In Figure 4c, participant B dies before the 36 month landmark time, and is not included in the model. Participant A shows a slight decrease in Fine and Gross Motor, while Speech & Swallow is similar, resulting in no steep drops in their predicted survival probabilities. Participant C also shows no important decreases after 24 months in the subscores, resulting in no drastic changes in their predicted survival probabilities when compared with their prediction in Figure 4b. These results highlight the ability of our model to accurately obtain disease trajectories for individuals and dynamically predict survival probabilities as new clinical data is accrued.

### 3.4 External Validation

An external dataset provided by Emory University (“Emory data”) was used to quantify the predictive capabilities of our model. The Emory data comes from the Emory ALS Clinic database, which collects clinical and demographic variables from patient visits since 1997^35^. The ALS population from the Emory data are predominantly from the state of Georgia, and thus do not overlap with the geographic regions encompassed by the ALS NHC. This provides an opportunity to verify the predictive capabilities of our model in a different population.

Web Table 1 presents the main characteristics of the data from Emory University’s ALS cohort compared to the NHC cohort. We used the Standardized Mean Difference (SMD) ^36,37^ to compare the distribution of variables between both cohorts. Both cohorts have slight differences on all variables except gender, which presents no difference. Riluzole use shows a very large difference between both cohorts. Adherence to riluzole use is variable in ALS populations ^38,39^. Given the length of time from which Emory’s observations were recorded, difference in riluzole use was not deemed a problem.

The fitted model on the NHC data was used to predict survival probabilities for participants in the Emory data at each of the landmark times. Additionally, a non-functional landmarking model was fit to compare its predictive capabilities against the FCLM. This was defined as a Cox model fit using the last observed ALSFRS-R values for each participant in the corresponding landmark timeframe as a scalar covariate. The integrated Brier Score (IBS) ^40^ and Harrell’s C-Index ^41^ were used to measure the predictive capabilities of both models. The IBS is obtained by integrating the Brier Score (BS) at predefined timepoints, with lower values indicating better predictive performance. In this case, at 0, 6, 12, 18, and 24 months the BS for each model was calculated and numerically integrated to obtain the IBS. Harrell’s C-Index quantifies the probability that, out of two randomly chosen patients, the one with a higher risk 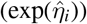 will have a shorter survival time than the one with a lower risk, with higher values indicating better performance capabilities. Both metrics can take values between 0 and 1, with a value of 0.25 or lower being used as a rule of thumb for good predictive capabilities for the IBS, and a value of 0.5 for the C-Index indicating that the model performs as well as a coin-flip. Table 2 presents the IBS and C-Indices for the FCLM model compared to a non-functional landmarking model when predicting for the Emory dataset.

**TABLE 2.** IBS and C-Index for a non-functional landmarking model and the FCLM, from the Emory external validation data, at landmark times of 12, 24, and 36 months.

| **Landmark Time** | **Landmark** |  | **FCLM** |  |
| --- | --- | --- | --- | --- |
|  | **IBS** | **C-Index** | **IBS** | **C-Index** |
| 12 months | 0.155 | 0.705 | **0.152** | **0.718** |
| 24 months | 0.168 | 0.662 | **0.156** | **0.712** |
| 36 months | 0.165 | 0.657 | **0.155** | **0.738** |

The FCLM has lower IBS values and higher C-Indices across all landmark times when compared to the non-functional landmark model, with the differences being quite stark for later landmark times. Across landmark times, the FCLM shows consistent values for both IBS and C-Indices. In contrast, the non-functional model performs worse at later landmark times. These results showcase the replicability of the predictive capabilities of our model to a different dataset. The Emory cohort came from a different region and was observed over a longer period of time than the ALS NHC cohort. The fact that our model gave consistently high predictive results for the external validation in the Emory data highlights both the flexibility of and consistency in predictions from our model when faced with new data.

## 4 SIMULATION STUDY

For our simulation study, we consider the case with only one functional covariate *W*_*i*_ and no scalar covariates at one specific landmark time. The model introduced in (3) can be simplified to log 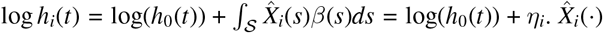 are the predicted individual trajectory curves from the sparse FPCA described in Subsection 2.2.

Following the procedure of Cui et al. ^22^, a summary of the steps used in our simulation procedure is described as follows: 1. Obtain an estimate of the cumulative baseline hazard function 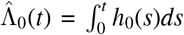. 2. Simulate individual trajectory curves 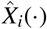 based on the sparse FPCA used to fit the model. 3. Define the functional coefficients *β*(*·*) to obtain 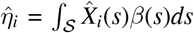.4. Estimate the survival function as 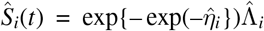. 5. Simulate the survival times 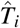 from *Ŝ*_*i*_(*t*). 6. Sample the censoring time *Ĉ*_*i*_ from the original data. 7. Derive the simulated observed survival time 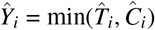. Using this procedure, we can simulate both predicted individual trajectories from a sparse FPCA, as well as survival times using specific forms of the functional coefficient *β*(*·*).

We used results from a model fit using the total ALSFRS-R score from the ALS NHC dataset at the 12 month landmark, which is described in Subsection 3.1, to obtain the cumulative baseline hazard function 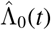. Using this model, we obtain the estimated cumulative baseline hazard and use a shape constrained smoother to ensure non-decreasing and non-negative values over the whole functional domain. For the simulation of 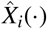, we simulate the FPCA scores for each individual, across all *K* principal components, such that 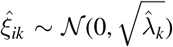. Once we simulate all the FPCA scores, we use (2) to directly simulate the predicted individual trajectories, using the estimated mean function 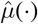 and eigen-functions 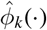 from the fitted model. We defined two general cases for *β*(*·*): based on real data, or following certain functions of interest. The details of the different scenarios defined for *β*(*s*) are presented in Section 4.1.

### 4.1 Scenarios

For this simulation study, special interest lies in comparing the performance of the model to estimate different possible forms for the functional coefficient *β*(*·*). Two forms of defining *β*(*·*) will be done: based on the functional form obtained from the ALS NHC dataset (“Real Data”), and using pre-specified function (“Pre-specified”). For the Real Data scenarios, a FCLM was fit at the 12, 24, and 36 month landmark times, and the estimated functional coefficient *β*(*·*) for each one was taken. For comparison purposes, the ALSFRS-R scores were scaled to be between 0 and 1.

For the Pre-specified scenarios, we define a series of functional forms for *β*(*·*) to account for different characteristics that might be observed in practice. These functional forms are:

1. 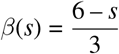;
2. 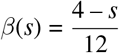;
3. 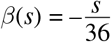;
4. 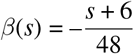;
5. 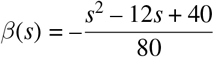;
6. 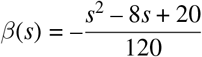;
7. 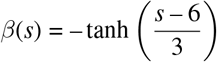;
8. 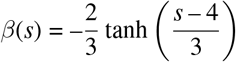;
9. 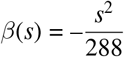;and
10. 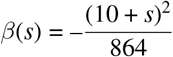;

A series of constants were added to the functional forms to ensure linear predictors, and thus simulated survival times, that were in line with the observed data.

Two broad groups of functions were defined, with scenarios I-IV corresponding to linear functions, while V-X are nonlinear. For scenarios I-X, the model being used was fitted at the landmark time of 12 months so *s* = [0, 12]. Thus the functional coefficients in scenarios I, V, and VII are symmetric, while all other scenarios are asymmetric. The functional coefficients in scenarios IV, V, VI, and X have no roots, for scenarios V and VI are non-monotonic, and for scenarios VII and VIII are concave.

100 simulated datasets were generated for each scenario and the fitted coefficients from the linear functional Cox model were obtained. Apart from fitting the linear functional Cox landmarking model, for each simulated dataset the last simulated value of the functional variable was used to fit a non-functional Cox landmark model for comparison purposes.

### 4.2 Performance Metrics

To measure our model’s capabilities to estimate different functional coefficient shapes the integrated square error (ISE) was used. The ISE is defined as 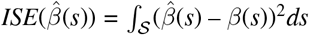, where *β*(*s*) is the known real functional coefficient and 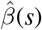 sis the estimate obtained from the model. Since the non-functional landmark model only estimates the coefficient at one time-point the ISE can not be calculated and thus was not included. To measure predictive performance both the IBS and C-Index were used. These measures are further described in Subsection 3.4.

### 4.3 Results

Table 3 presents the median MSE/ISE, IBS, and C-index, for each simulation scenario, with bolded numbers for each metric representing which model had better performance.

**TABLE 3.**
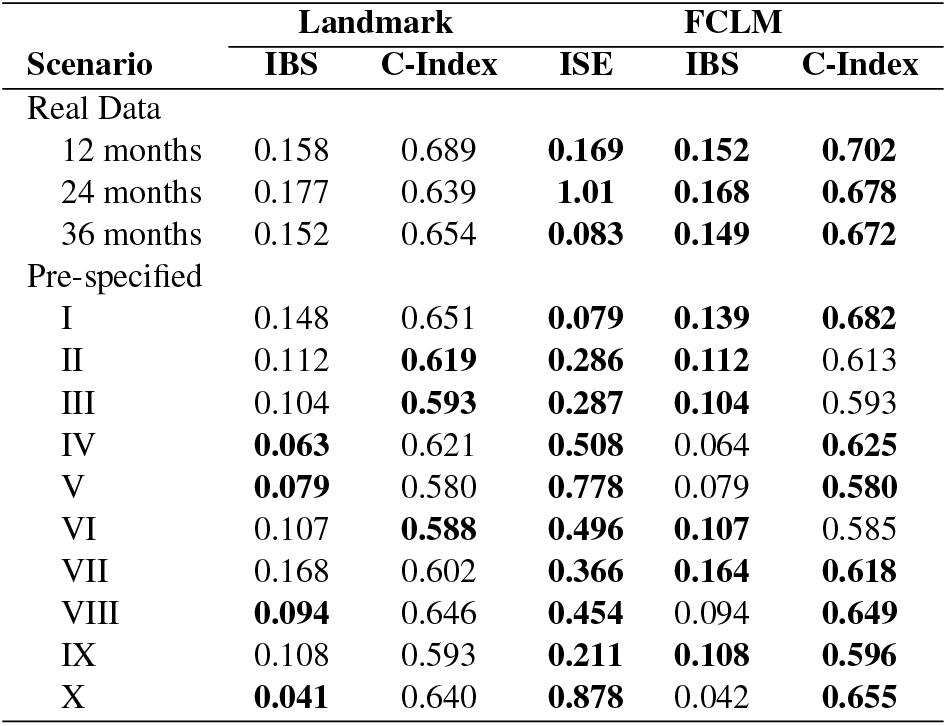
Median IBS, C-Index, and ISE, for each of the simulation scenarios, by model used for estimation.

Our model presents good performance metrics overall, with small ISEs for most simulation scenarios, and better, or equal, performance metrics for almost all scenarios. The results are good when estimating the functional coefficient from the Real Data scenarios, with most simulation scenarios being comparable based on the ISE. The simulated functional coefficients from the Real Data scenarios for 12 and 36 months had some of the lowest median ISEs, while Pre-specified scenarios V, X, and Real Data scenario for 24 months had the highest values. The median ISE for the FCLM was considerably lower in all scenarios than the MSE for the non-functional landmarking model.

In terms of predictive capabilities, the Real Data scenarios had relatively high C-indices and low IBS, which were better than those from a landmark model using only the last observed value of the covariate. For the Pre-specified scenarios in most scenarios the IBS was as good as the one from the non-functional landmark model, with some scenarios showing a slight improvement in this metric. The improvements are more evident in the C-indices, with most scenarios having an increase in this metric. For a variety of different scenarios, both across landmark times and with different functional coefficients, our model shows better performance when compared to a non-functional landmark model.

## 5 DISCUSSION

Our proposed landmarking model allows for dynamic prediction of survival probabilities based on changes to sparse multivariate functional variables. Estimating the underlying disease progression process allows for a more detailed analysis of disease progression when compared to using a simple summary statistic. The use of a functional covariate permits the estimation of a functional coefficient, which describes the relationship between the sparse functional covariates over time with respect to the time-to-event outcome. Specific trends in disease progression can be studied and leveraged to have more accurate predictions of time to important disease progression outcomes.

When analyzing the NHC data, the strengths of the two-steps of our approach can be seen. The multivariate sparse FPCA can generate principal components which discriminate between the subscores, while at the same time obtaining estimates for not only the correlation within subscores, but also across subscores. When including the predicted individual trajectories from the first step into the landmarking model, we can obtain different functional coefficients for each subscore. The dynamic prediction aspect of the landmarking model can be useful for easily predicting survival curves for patients at different times of interest, based on their disease progression history. This two-step approach allows the prediction of both the estimated individual trajectory curves as well as the survival probabilities to be updated as new disease progression information is accrued.

The external validation using Emory’s ALS dataset shows the robustness of out-of-sample predictions from this model, and the stability in predictive metrics across landmark times. For a more detailed application of non-functional landmark models on time-to-event outcomes within the context of ALS, see Schneck et al. ^42^.

The simulation results show the capabilities of the linear functional Cox model to recover different forms of the functional coefficient. We highlight that our model can recover functional forms that did not have roots or were non-linear.

Although our modeling approach extends the landmarking framework to include sparse multivariate functional data, there are still additional topics to explore. The ALSFRS-R subscores are bounded by construction to take values between 0 and Techniques to address this in the modelling approach were not analyzed due to adding several degrees of complication. The selection of the landmarking times is an important topic that requires further study. In a clinical setting patients might be enrolled well after their initial diagnosis, excluding them from several landmark times. Further extensions to accomodate for this staggered enrollment or left-truncated data are of interest.

## Data Availability

The data that support the findings of this study are available from the corresponding author upon reasonable request.

## ACKNOWLEDGMENTS

The authors would like to thank Dr. Jonathan Glass and Dr. Christina Fournier from the Emory ALS Center for providing the data used for external validation.

## SUPPORTING INFORMATION

Additional supporting information can be found online in the Supporting Information section.

