## Supplemental Material for "Sparse Multivariate Functional Cox Landmarking Model with Applications to ALS Progression"

Web Table 1: Demographic and clinical characteristics of participants, included in the analytic sample, from Emory University's ALS data compared to those from the NHC data. Summary statistics are: median (IQR) for numerical variables and n (%) for categorical variables. Standardized mean differences (SMD) are presented for comparisons.

| **Characteristic** | **Emory** (N = 1,277) | **NHC** (N = 1,482) | **SMD** |
|---|---|---|---|
| Male | 753 (59.0%) | 874 (59.0%) | 0.00 |
| Limb onset site | 937 (73.4%) | 1,001 (67.5%) | 0.13 |
| Age at Onset (years) | 60.0 (51.0, 68.3) | 62.4 (54.7, 69.2) | 0.21 |
| Age at Diagnosis (years) | 61.60 (53.1, 69.5) | 64.0 (56.0, 71.0) | 0.20 |
| Uses Riluzole | 441 (34.5%) | 1,172 (79.1%) | 1.0 |
| Number of ALSFRS visits | 3 (1, 5) | 4 (3, 7) | 0.24 |

Web Figure 1: Estimated functional coefficient $\hat{\beta}(s)$ with 95% confidence band, for each of the ALSFRS-R subscores, across time, at the two year landmark

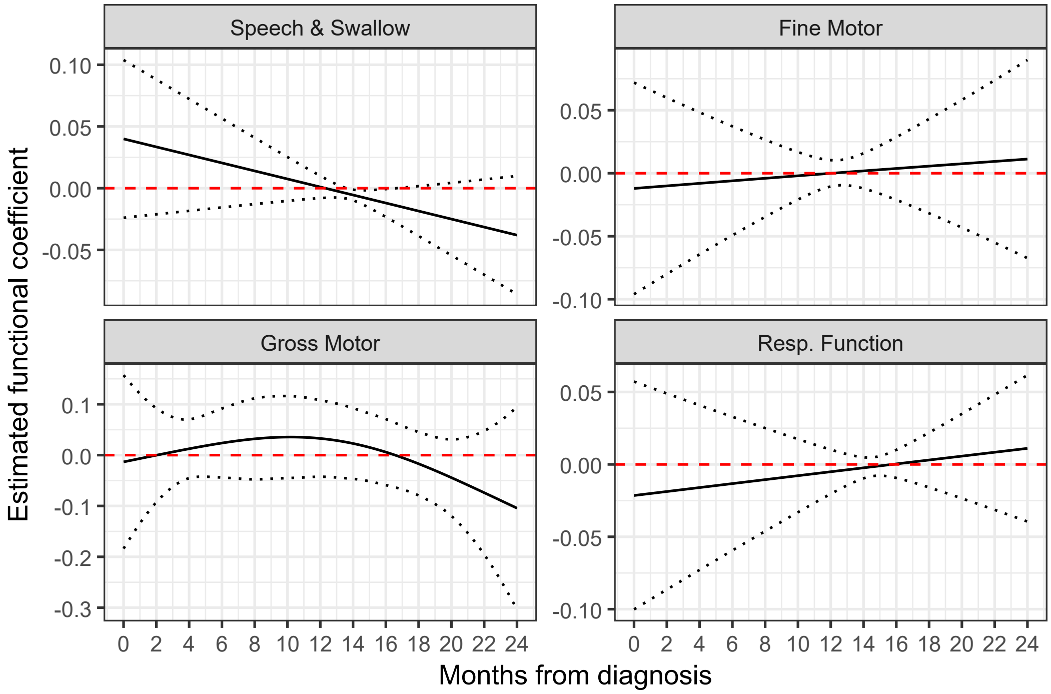

Web Figure 2: Estimated functional coefficient $\hat{\beta}(s)$ with 95% confidence band, for each of the ALSFRS-R subscores, across time, at the three year landmark

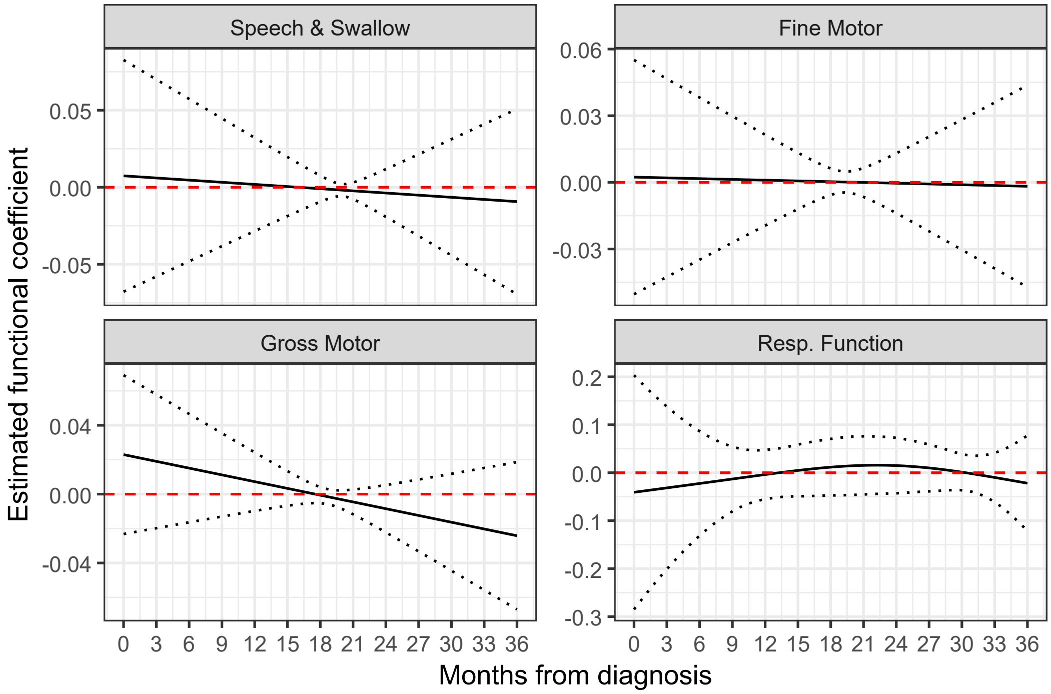
